# Vaccines Elicit Highly Cross-Reactive Cellular Immunity to the SARS-CoV-2 Omicron Variant

**DOI:** 10.1101/2022.01.02.22268634

**Authors:** Jinyan Liu, Abishek Chandrashekar, Daniel Sellers, Julia Barrett, Michelle Lifton, Katherine McMahan, Michaela Sciacca, Haley VanWyk, Cindy Wu, Jingyou Yu, Ai-ris Y. Collier, Dan H. Barouch

## Abstract

The highly mutated SARS-CoV-2 Omicron (B.1.1.529) variant has been shown to evade a substantial fraction of neutralizing antibody responses elicited by current vaccines that encode the WA1/2020 Spike immunogen^1^, resulting in increased breakthrough infections and reduced vaccine efficacy. Cellular immune responses, particularly CD8+ T cell responses, are likely critical for protection against severe SARS-CoV-2 disease^2-6^. Here we show that cellular immunity induced by current SARS-CoV-2 vaccines is highly cross-reactive against the SARS-CoV-2 Omicron variant. Individuals who received Ad26.COV2.S or BNT162b2 vaccines demonstrated durable CD8+ and CD4+ T cell responses that showed extensive cross-reactivity against both the Delta and Omicron variants, including in central and effector memory cellular subpopulations. Median Omicron-specific CD8+ T cell responses were 82-84% of WA1/2020-specific CD8+ T cell responses. These data suggest that current vaccines may provide considerable protection against severe disease with the SARS-CoV-2 Omicron variant despite the substantial reduction of neutralizing antibody responses.

---

Recent studies have shown that vaccine-elicited neutralizing antibodies (NAbs) are substantially reduced to the highly mutated SARS-CoV-2 Omicron variant, resulting in rapid global spread, including breakthrough infections in fully vaccinated individuals^1^. To evaluate the cross-reactivity of vaccine-elicited cellular immune responses against the SARS-CoV-2 Omicron variant, we assessed CD8+ and CD4+ T cell responses in 51 individuals who were vaccinated with the adenovirus vector-based Ad26.COV2.S vaccine^7^ (Johnson & Johnson; N=20) or the mRNA-based BNT162b2 vaccine^8^ (Pfizer; N=31).

Following BNT162b2 vaccination, we observed high WA1/2020-specific NAb responses at month 1, followed by a sharp decline by month 8, as expected^9,10^ (**Fig. 1a**). Following Ad26.COV2.S vaccination, there were substantially initial lower WA1/2020-specific NAb responses at month 1, but these responses were more durable and persisted at month 8^9,11^ (**Fig. 1a**). However, minimal cross-reactive Omicron-specific NAbs were observed for both vaccines (**Fig. 1a**), consistent with recent data in the absence of additional boosting^1^. Receptor binding domain (RBD)-specific binding antibody responses were assessed by ELISA and showed similar trends, with minimal cross-reactive Omicron-specific binding antibodies (**Fig. 1b**).

**Figure 1.**
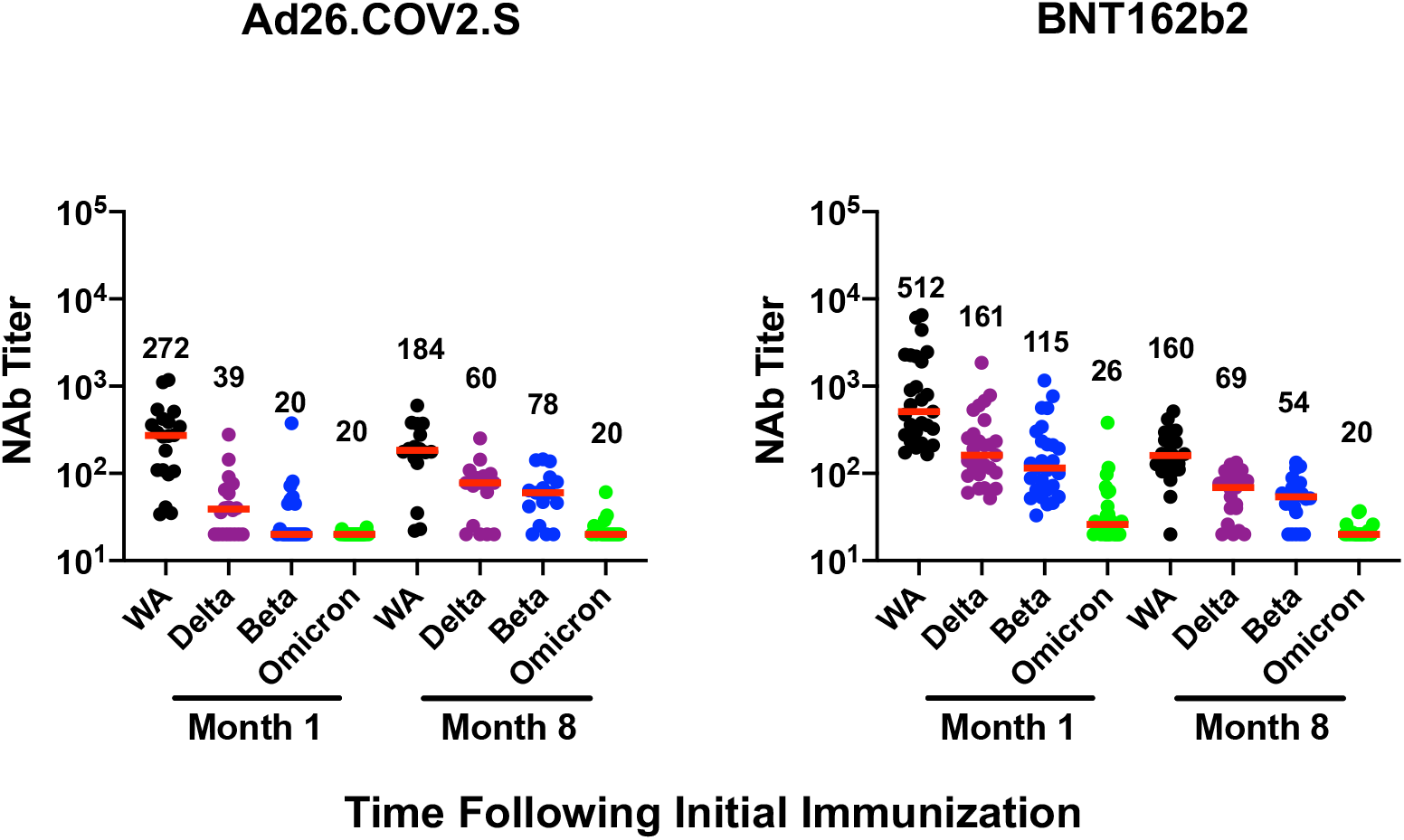
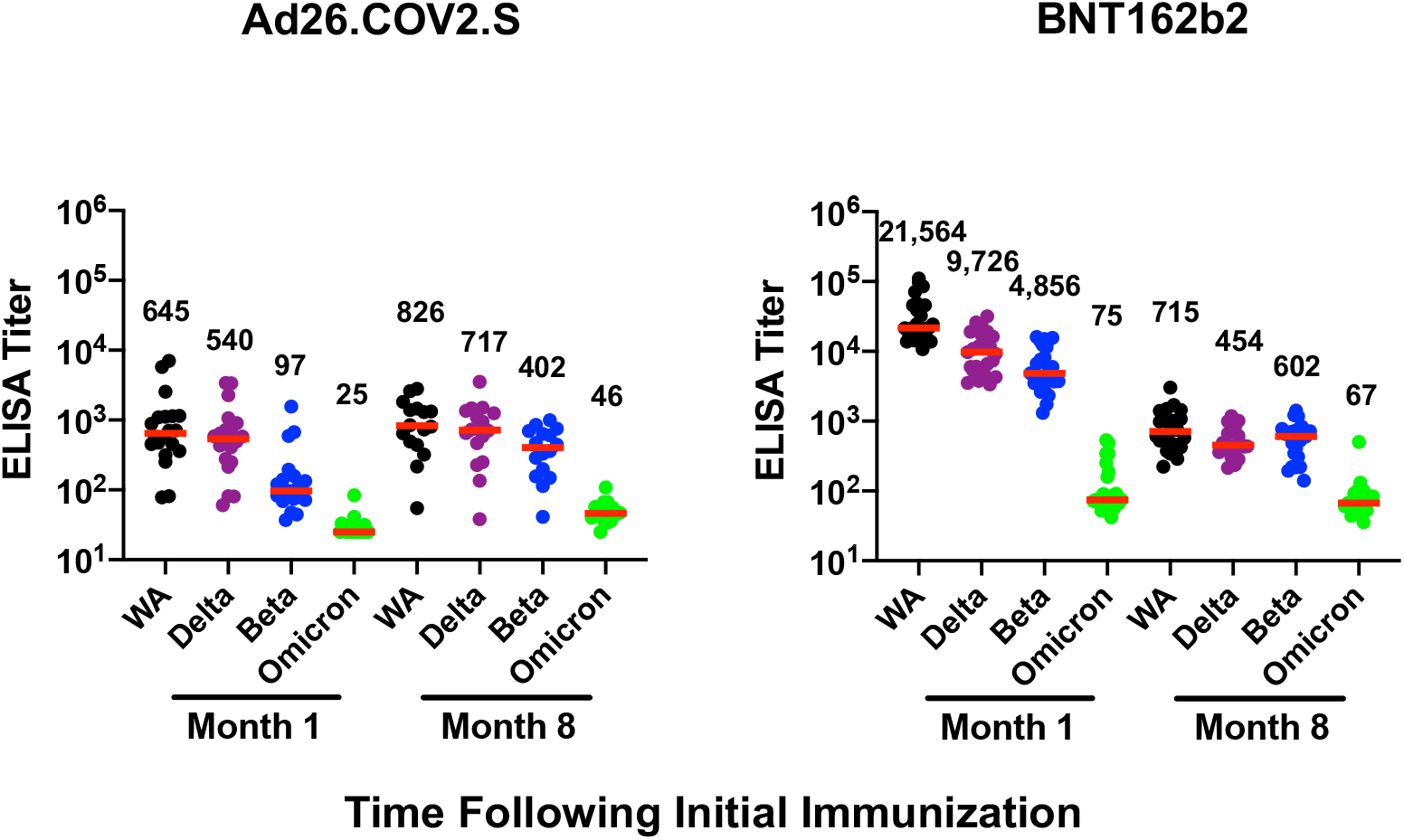
Humoral immune responses to Omicron. Antibody responses at months 1 and 8 following vaccination with Ad26.COV2.S or BNT162b2. **a**, Neutralizing antibody (NAb) titers by a luciferase-based pseudovirus neutralization assay. **b**, Receptor binding domain (RBD)-specific binding antibody titers by ELISA. Responses were measured against the SARS-CoV-2 WA1/2020, B.1.617.2 (Delta), B.1.351 (Beta), and B.1.1.529 (Omicron) variants. Medians (red bars) are depicted and numerically shown.

In contrast with antibody responses, Spike-specific cellular immune responses assessed by pooled peptide IFN-γ ELISPOT assays showed substantial cross-reactivity to Omicron (**Extended Data Fig. 1**). We next assessed Spike-specific CD8+ and CD4+ T cell responses by intracellular cytokine staining assays. Ad26.COV2.S induced median Spike-specific IFN-γ CD8+ T cell responses of 0.061%, 0.062%, and 0.051% against WA1/2020, Delta, and Omicron, respectively, at month 8 following vaccination (**Fig. 2a**). BNT162b2 induced median Spike-specific IFN-γ CD8+ T cell responses of 0.028% and 0.023% against WA1/2020 and Omicron, respectively, at month 8 following vaccination (**Fig. 2a**). These data suggest that Omicron-specific CD8+ T cell responses were 82-84% cross-reactive with WA1/2020-specific CD8+ T cell responses. Spike-specific IFN-γ CD4+ T cell responses elicited by Ad26.COV2.S were a median of 0.026%, 0.030%, and 0.029% against WA1/2020, Delta, and Omicron, respectively, and by BNT162b2 were a median of 0.033% and 0.027% against WA1/2020 and Omicron, respectively, indicating substantial cross-reactivity of CD4+ T cell responses as well (**Fig. 2b**). Substantial Omicron cross-reactivity was also observed for Spike-specific TNF-*α* and IL-2 secreting CD8+ and CD4+ T cell responses (**Extended Data Fig. 2**).

**Figure 2.**
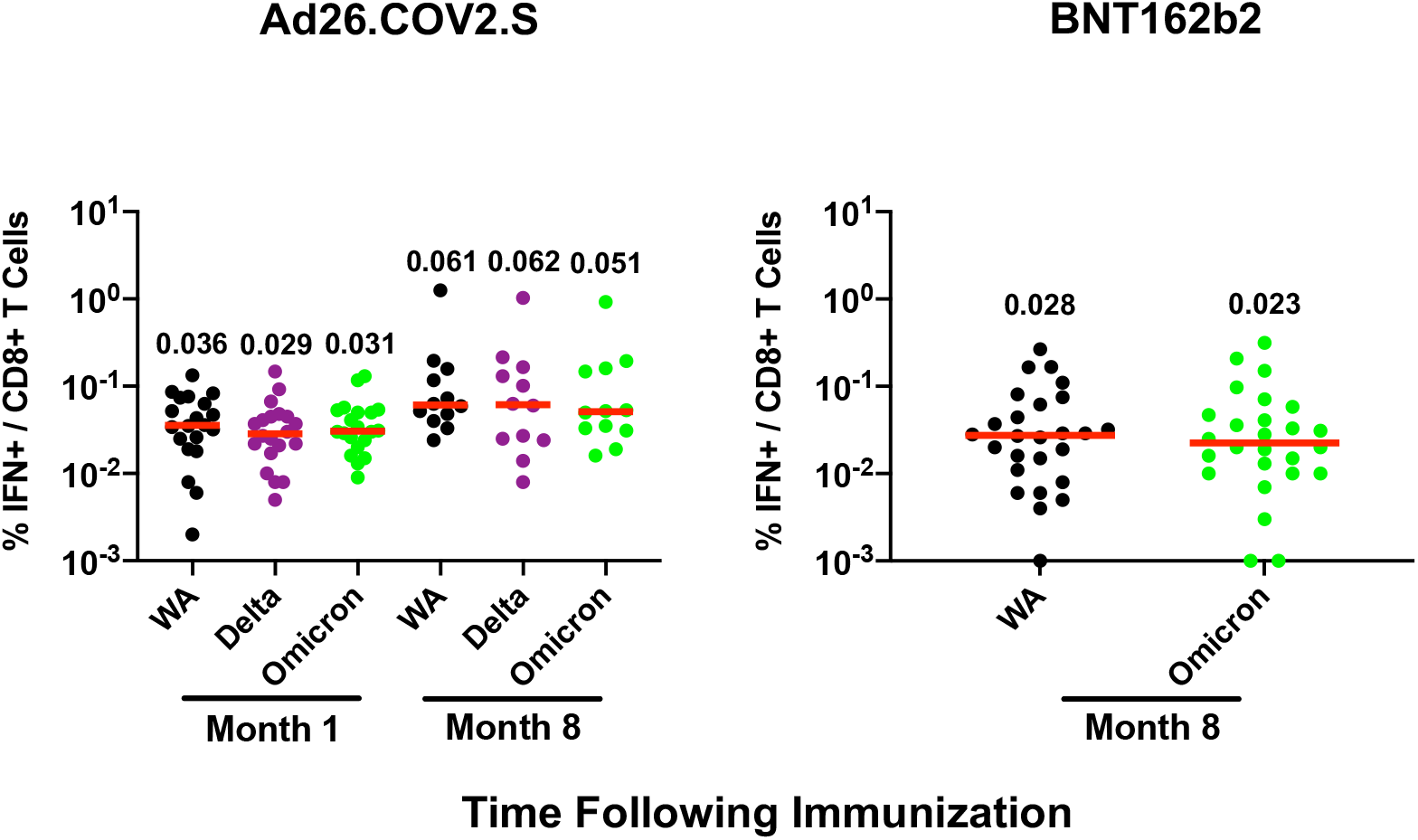
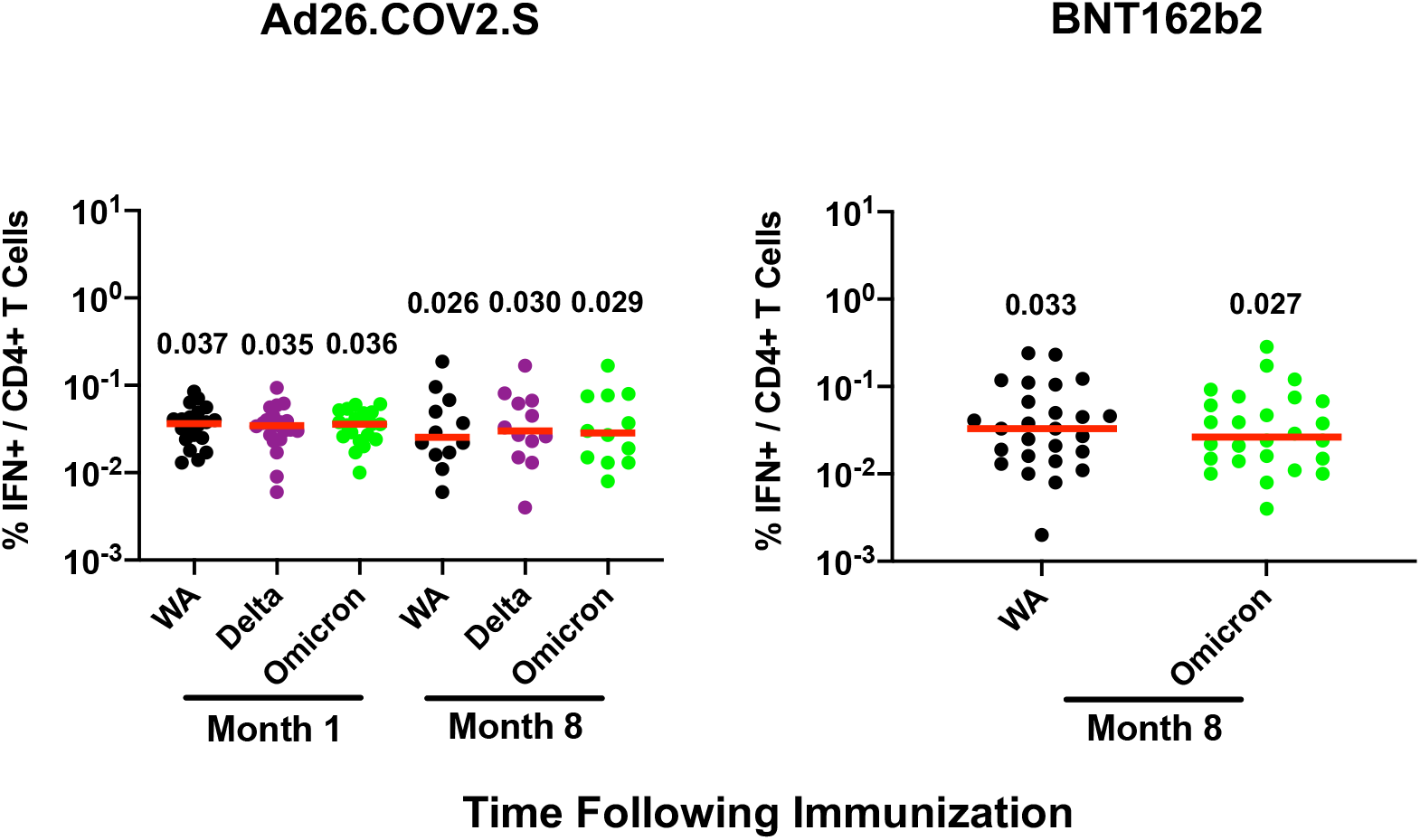
Cellular immune responses to Omicron. T cell responses at months 1 and 8 following vaccination with Ad26.COV2.S or BNT162b2. Pooled peptide Spike-specific IFN-γ (**a**) CD8+ T cell responses and (**b**) CD4+ T cell responses by intracellular cytokine staining assays. Responses were measured against the SARS-CoV-2 WA1/2020, B.1.617.2 (Delta), and B.1.1.529 (Omicron) variants. Medians (red bars) are depicted and numerically shown.

Linear regression analysis showed that Omicron-specific CD8+ T cell responses correlated with WA1/2020-specific CD8+ T cell responses for the Ad26.COV2.S vaccine for both timepoints (R=0.78, P<0.0001, slope 0.75) and the BNT162b2 vaccine (R=0.56, P<0.0001, slope 0.81), although two individuals had undetectable Omicron-specific CD8+ T cell responses following BNT162b2 vaccination (**Fig. 3a**). Similarly, Omicron-specific CD4+ T cell responses correlated with WA1/2020-specific CD4+ T cell responses for both the Ad26.COV2.S vaccine (R=0.79, P<0.0001, slope 0.83) and the BNT162b2 vaccine (R=0.90, P<0.0001, slope 0.88) (**Fig. 3b**).

**Figure 3.**
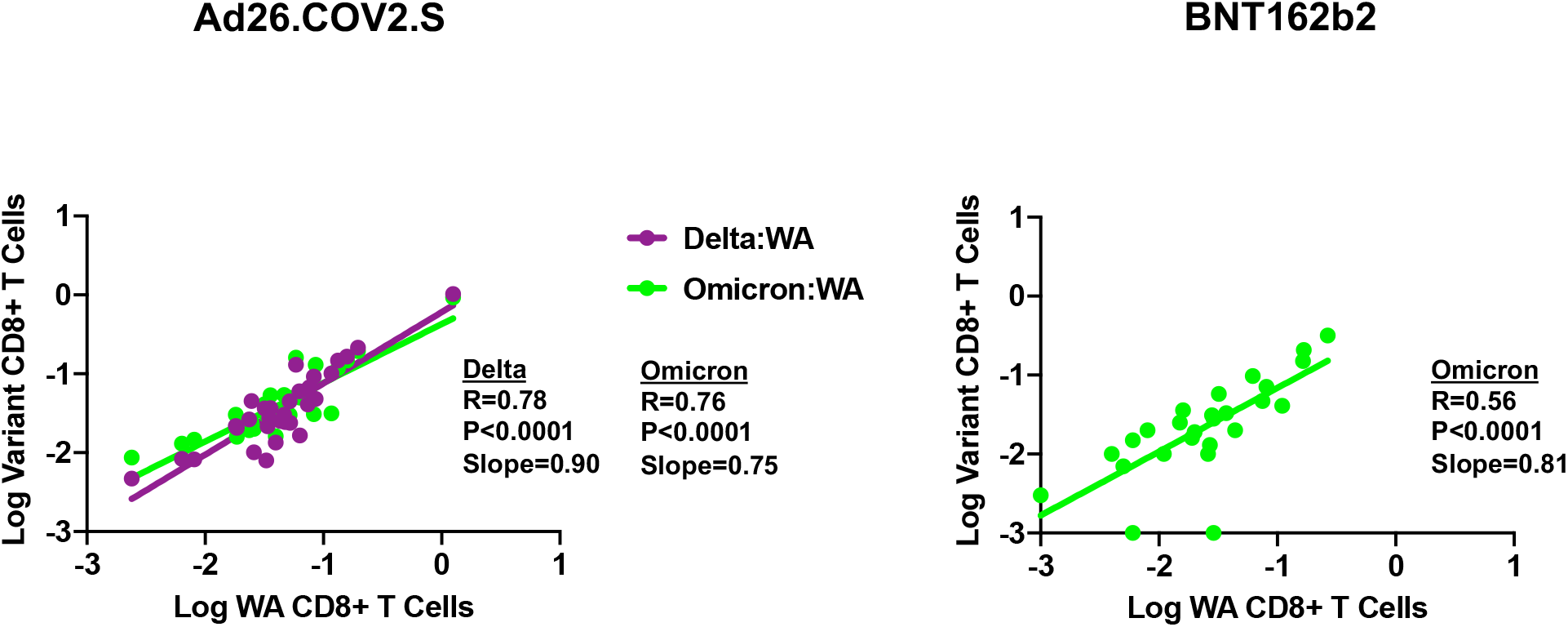
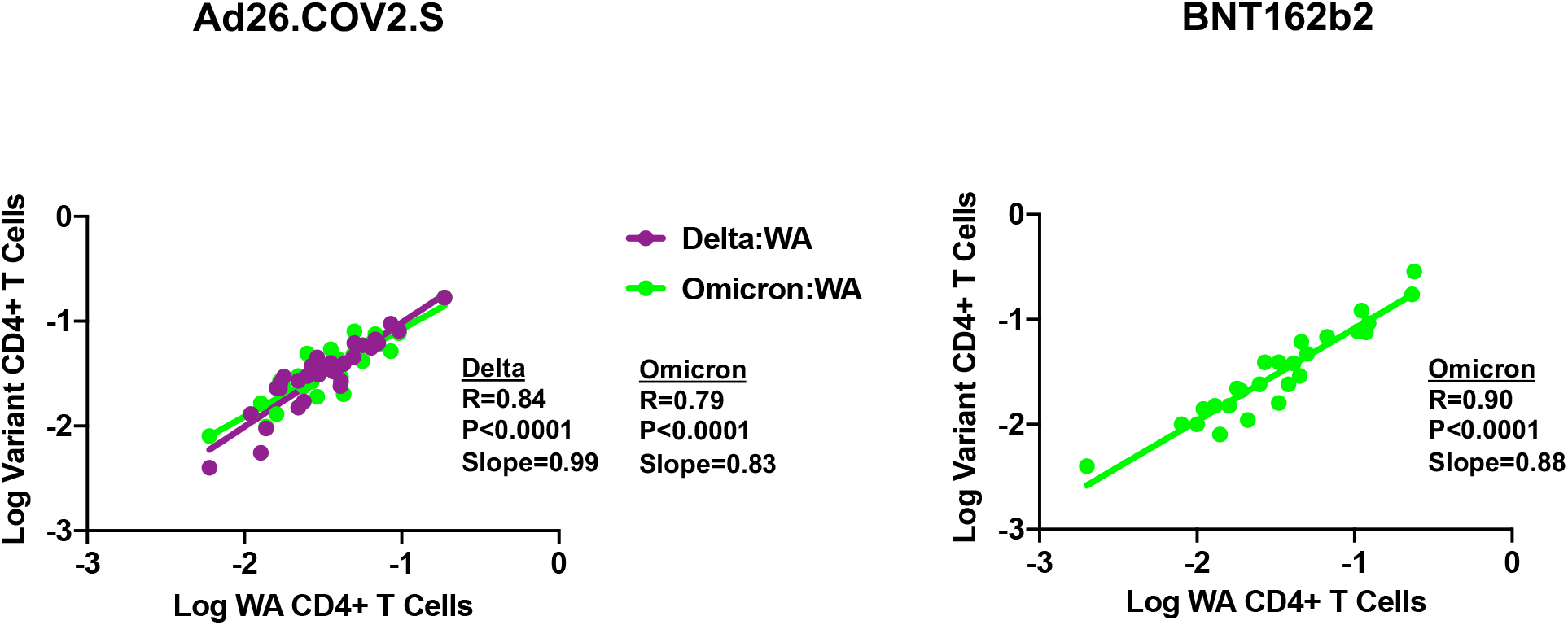
Correlations of variant- and WA1/2020-specific cellular immune responses. Correlations of Log Delta- and Omicron-specific to Log WA1/2020-specific (**a**) CD8+ T cell responses and (**b**) CD4+ T cell responses by intracellular cytokine staining assays. Lines of best fit by logistic regression are shown.

Spike-specific IFN-γ CD8+ and CD4+ T cell central memory and effector memory subpopulations elicited by Ad26.COV2.S also showed extensive cross-reactivity to Delta and Omicron. At month 8, CD8+ central memory responses were 0.076%, 0.054%, and 0.075%, CD8+ effector memory responses were 0.168%, 0.143%, and 0.146%, CD4+ central memory responses were 0.030%, 0.035%, and 0.038%, and CD4+ effector memory responses were 0.102%, 0.094%, and 0.083%, against WA1/202, Delta, and Omicron, respectively (**Fig. 4**).

**Figure 4.**
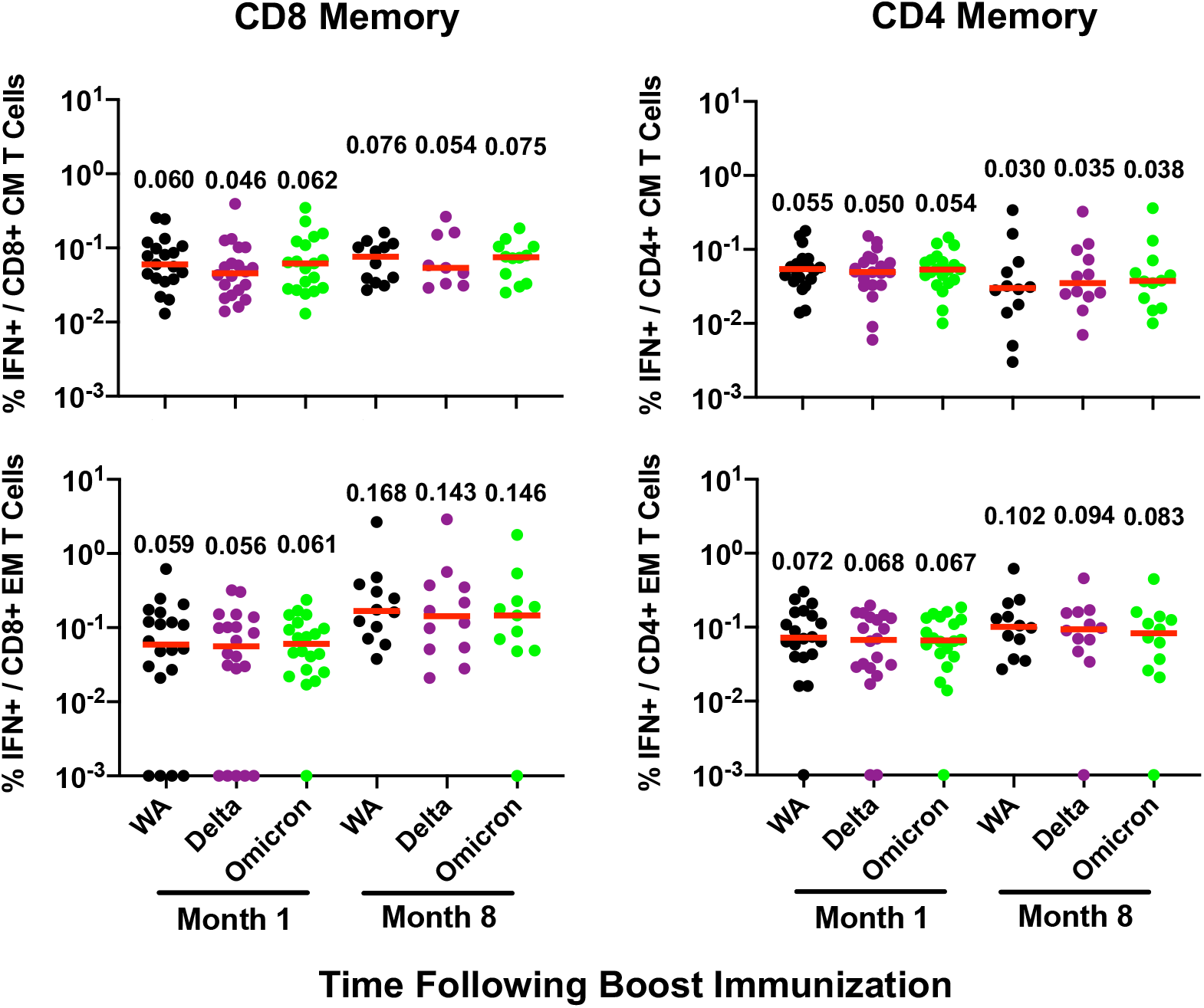
Cellular immune memory subpopulations to Omicron. Pooled peptide Spike-specific IFN-γ CD8+ and CD4+ central memory (CD45RA-CD27+) and effector memory (CD45RA-CD27-) T cell responses by intracellular cytokine staining assays at months 1 and 8 following vaccination with Ad26.COV2.S. Responses were measured against the SARS-CoV-2 WA1/2020, B.1.617.2 (Delta), and B.1.1.529 (Omicron) variants. Medians (red bars) are depicted and numerically shown.

Our data demonstrate that Ad26.COV2.S and BNT162b2 elicit broadly cross-reactive cellular immunity against SARS-CoV-2 variants including Omicron. The consistency of these observations across two different vaccine platform technologies (viral vector and mRNA) suggests the generalizability of these findings. The extensive cross-reactivity of Omicron-specific CD8+ and CD4+ T cell responses contrasts sharply with the markedly reduced Omicron-specific neutralizing and binding antibody responses. These data are consistent with prior studies showing greater cross-reactivity of vaccine-elicited cellular immune responses compared with humoral immune responses against the SARS-CoV-2 Alpha, Beta, and Gamma variants^12^. The 82-84% cross-reactivity of CD8+ T cell responses to Omicron is consistent with theoretical predictions based on the Omicron mutations^6^.

Preclinical studies have shown that CD8+ T cells contribute to protection against SARS-CoV-2 in rhesus macaques, particularly when antibody responses are suboptimal^5^. Durable CD8+ and CD4+ T cell responses have also been reported following infection and vaccination^2-4,6,9,11,13,14^. Given the role of CD8+ T cells in clearance of viral infections, it is likely that cellular immunity contributes substantially to vaccine protection against severe SARS-CoV-2 disease. This may be particularly relevant for Omicron, which evades a substantial portion of antibody responses. Our data suggest that current vaccines may provide substantial protection against severe disease due to the SARS-CoV-2 Omicron variant despite reduced neutralizing antibody responses and increased breakthrough infections.

## Data Availability

All data produced in the present work are contained in the manuscript

## Acknowledgements

The authors acknowledge NIH grant CA260476, the Massachusetts Consortium for Pathogen Readiness, the Ragon Institute, and the Musk Foundation (D.H.B.) as well as the Reproductive Scientist Development Program from the Eunice Kennedy Shriver National Institute of Child Health & Human Development and Burroughs Wellcome Fund HD000849 (A.Y.C.).

## Data sharing

All data are available in the manuscript or the supplementary material. Correspondence and requests for materials should be addressed to D.H.B.

## Author Contributions

This study was designed by DHB. Samples were provided by AYC. Cellular immune responses were assessed by JL, AC, DS, JB, ML, MS, HV, and CW. Humoral immune responses were assessed by KM and JY.

## Methods

### Study population

Samples from individuals who received the BNT162b2 vaccine were obtained from the Beth Israel Deaconess Medical Center (BIDMC) specimen biorepository. Samples from individuals who received Ad26.COV2.S were obtained from the COV1001 study (NCT04436276). Both studies were approved by the BIDMC institutional review board. All participants provided informed consent. Individuals were excluded from this study if they had a history of SARS-CoV-2 infection, received other COVID-19 vaccines, or received immunosuppressive medications.

### Pseudovirus neutralizing antibody assay

The SARS-CoV-2 pseudoviruses expressing a luciferase reporter gene were used to measure pseudovirus neutralizing antibodies. In brief, the packaging construct psPAX2 (AIDS Resource and Reagent Program), luciferase reporter plasmid pLenti-CMV Puro-Luc (Addgene) and spike protein expressing pcDNA3.1-SARS-CoV-2 SΔCT were co-transfected into HEK293T cells (ATCC CRL_3216) with lipofectamine 2000 (ThermoFisher Scientific). Pseudoviruses of SARS-CoV-2 variants were generated by using WA1/2020 strain (Wuhan/WIV04/2019, GISAID accession ID: EPI_ISL_402124), B.1.1.7 variant (Alpha, GISAID accession ID: EPI_ISL_601443), B.1.351 variant (Beta, GISAID accession ID: EPI_ISL_712096), B.1.617.2 (Delta, GISAID accession ID: EPI_ISL_2020950), or B.1.1.529 (Omicron, GISAID ID: EPI_ISL_7358094.2). The supernatants containing the pseudotype viruses were collected 48h after transfection; pseudotype viruses were purified by filtration with 0.45-μm filter. To determine the neutralization activity of human serum, HEK293T-hACE2 cells were seeded in 96-well tissue culture plates at a density of 1.75 × 10^4^ cells per well overnight. Three-fold serial dilutions of heat-inactivated serum samples were prepared and mixed with 50 μl of pseudovirus. The mixture was incubated at 37 °C for 1 h before adding to HEK293T-hACE2 cells. After 48 h, cells were lysed in Steady-Glo Luciferase Assay (Promega) according to the manufacturer’s instructions. SARS-CoV-2 neutralization titers were defined as the sample dilution at which a 50% reduction (NT50) in relative light units was observed relative to the average of the virus control wells.

### Enzyme-linked immunosorbent assay (ELISA)

SARS-CoV-2 spike receptor-binding domain (RBD)-specific binding antibodies in serum were assessed by ELISA. 96-well plates were coated with 2 μg/mL of SARS-CoV-2 WA1/2020, B.1.617.2 (Delta), B.1.351 (Beta), or B.1.1.529 (Omicron) RBD protein in 1× Dulbecco phosphate-buffered saline (DPBS) and incubated at 4 °C overnight. After incubation, plates were washed once with wash buffer (0.05% Tween 20 in 1× DPBS) and blocked with 350 μL of casein block solution per well for 2 to 3 hours at room temperature. Following incubation, block solution was discarded and plates were blotted dry. Serial dilutions of heat-inactivated serum diluted in Casein block were added to wells, and plates were incubated for 1 hour at room temperature, prior to 3 more washes and a 1-hour incubation with a 1:4000 dilution of anti– human IgG horseradish peroxidase (HRP) (Invitrogen, ThermoFisher Scientific) at room temperature in the dark. Plates were washed 3 times, and 100 μL of SeraCare KPL TMB SureBlue Start solution was added to each well; plate development was halted by adding 100 μL of SeraCare KPL TMB Stop solution per well. The absorbance at 450 nm, with a reference at 650 nm, was recorded with a VersaMax microplate reader (Molecular Devices). For each sample, the ELISA end point titer was calculated using a 4-parameter logistic curve fit to calculate the reciprocal serum dilution that yields a corrected absorbance value (450 nm-650 nm) of 0.2. Interpolated end point titers were reported.

### Enzyme-linked immunospot (ELISPOT) assay

ELISPOT plates were coated with mouse anti-human IFN-γ monoclonal antibody from MabTech at 1 µg/well and incubated overnight at 4°C. Plates were washed with DPBS, and blocked with R10 media (RPMI with 10% heat inactivated FBS with 1% of 100x penicillin-streptomycin, 1M HEPES, 100mM Sodium pyruvate, 200mM L-glutamine, and 0.1% of 55mM 2-Mercaptoethanol) for 2-4 h at 37°C. SARS-CoV-2 pooled S peptides from SARS-CoV-2 WA1/2020, B.1.617.2 (Delta), or B.1.1.529 (Omicron) (21st Century Biochemicals) were prepared and plated at a concentration of 2 µg/well, and 100,000 cells/well were added to the plate. The peptides and cells were incubated for 15-20 h at 37°C. All steps following this incubation were performed at room temperature. The plates were washed with ELISPOT wash buffer and incubated for 2-4 h with Biotinylated mouse anti-human IFN-γ monoclonal antibody from MabTech (1 µg/mL). The plates were washed a second time and incubated for 2-3 h with conjugated Goat anti-biotin AP from Rockland, Inc. (1.33 µg/mL). The final wash was followed by the addition of Nitor-blue Tetrazolium Chloride/5-bromo-4-chloro 3 ‘indolyphosphate p-toludine salt (NBT/BCIP chromagen) substrate solution for 7 min. The chromagen was discarded and the plates were washed with water and dried in a dim place for 24 h. Plates were scanned and counted on a Cellular Technologies Limited Immunospot Analyzer.

### Intracellular cytokine staining (ICS) assay

CD4+ and CD8+ T cell responses were quantitated by pooled peptide-stimulated intracellular cytokine staining (ICS) assays. Peptide pools contained 15 amino acid peptides overlapping by 11 amino acids spanning the SARS-CoV-2 WA1/2020, B.1.617.2 (Delta), or B.1.1.529 (Omicron) Spike proteins (21^st^ Century Biochemicals). 10^6^ peripheral blood mononuclear cells well were re-suspended in 100 µL of R10 media supplemented with CD49d monoclonal antibody (1 µg/mL) and CD28 monoclonal antibody (1 µg/mL). Each sample was assessed with mock (100 µL of R10 plus 0.5% DMSO; background control), peptides (2 µg/mL), and/or 10 pg/mL phorbol myristate acetate (PMA) and 1 µg/mL ionomycin (Sigma-Aldrich) (100µL; positive control) and incubated at 37°C for 1 h. After incubation, 0.25 µL of GolgiStop and 0.25 µL of GolgiPlug in 50 µL of R10 was added to each well and incubated at 37°C for 8 h and then held at 4°C overnight. The next day, the cells were washed twice with DPBS, stained with aqua live/dead dye for 10 mins and then stained with predetermined titers of monoclonal antibodies against CD279 (clone EH12.1, BB700), CD4 (clone L200, BV711), CD27 (clone M-T271, BUV563), CD8 (clone SK1, BUV805), CD45RA (clone 5H9, APC H7) for 30 min. Cells were then washed twice with 2% FBS/DPBS buffer and incubated for 15 min with 200 µL of BD CytoFix/CytoPerm Fixation/Permeabilization solution. Cells were washed twice with 1X Perm Wash buffer (BD Perm/WashTM Buffer 10X in the CytoFix/CytoPerm Fixation/ Permeabilization kit diluted with MilliQ water and passed through 0.22µm filter) and stained with intracellularly with monoclonal antibodies against IFN-γ (clone B27; BUV395), and CD3 (clone SP34.2, Alexa 700), for 30 min. Cells were washed twice with 1X Perm Wash buffer and fixed with 250µL of freshly prepared 1.5% formaldehyde. Fixed cells were transferred to 96-well round bottom plate and analyzed by BD FACSymphony(tm) system. Data were analyzed using FlowJo v9.9.

### Statistical analysis

Descriptive statistics and logistic regression were performed using GraphPad Prism 8.4.3, (GraphPad Software, San Diego, California).

## Extended Data Figure Legends

**Extended Data Figure 1.**
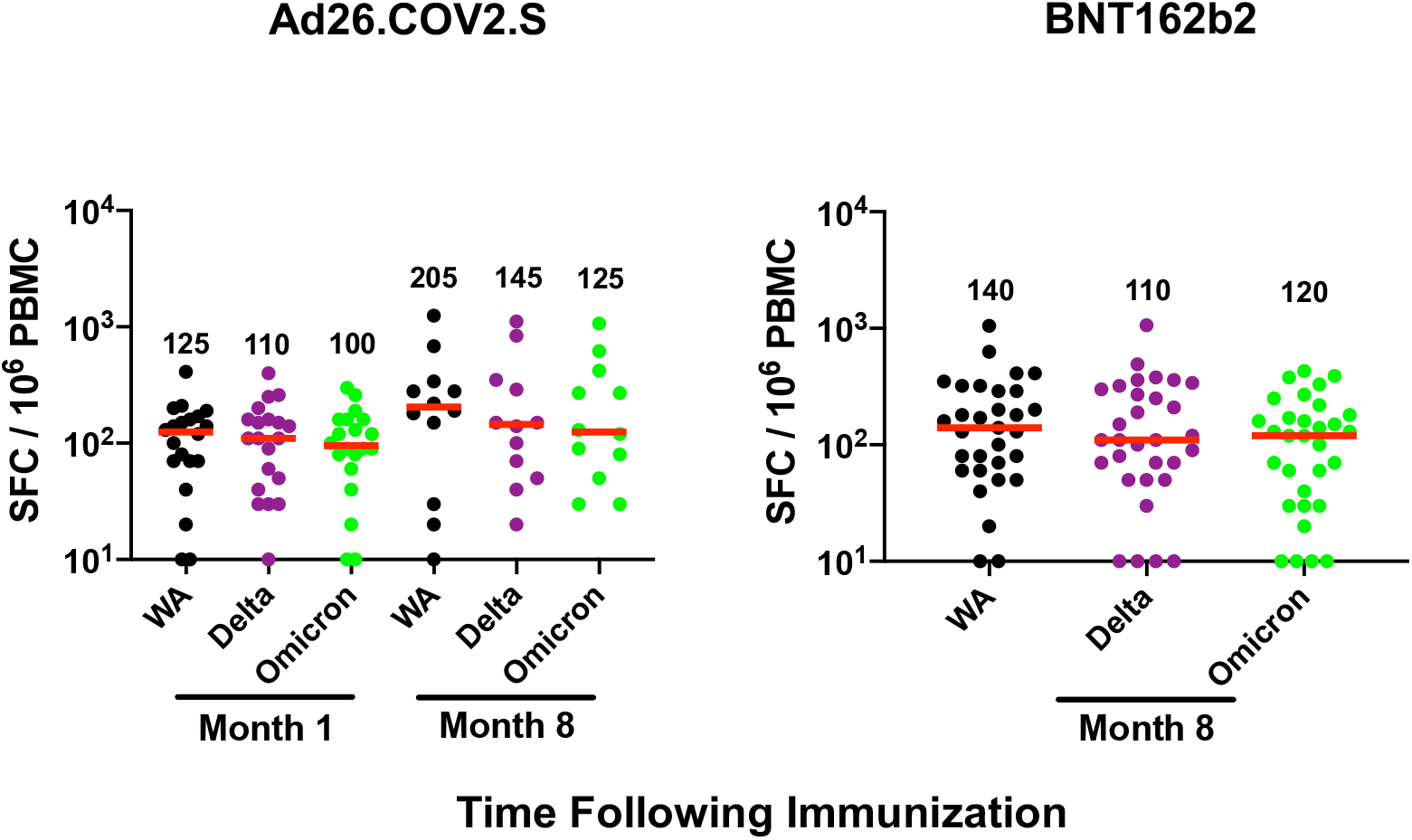
Cellular immune responses to Omicron by ELISPOT assays. Spike-specific IFN-γ ELISPOT assays at month 1 and 8 following vaccination with Ad26.COV2.S or BNT162b2. Responses were measured against the SARS-CoV-2 WA1/2020, B.1.617.2 (Delta), and B.1.1.529 (Omicron) variants. Medians (red bars) are depicted and numerically shown.

**Extended Data Figure 2.**
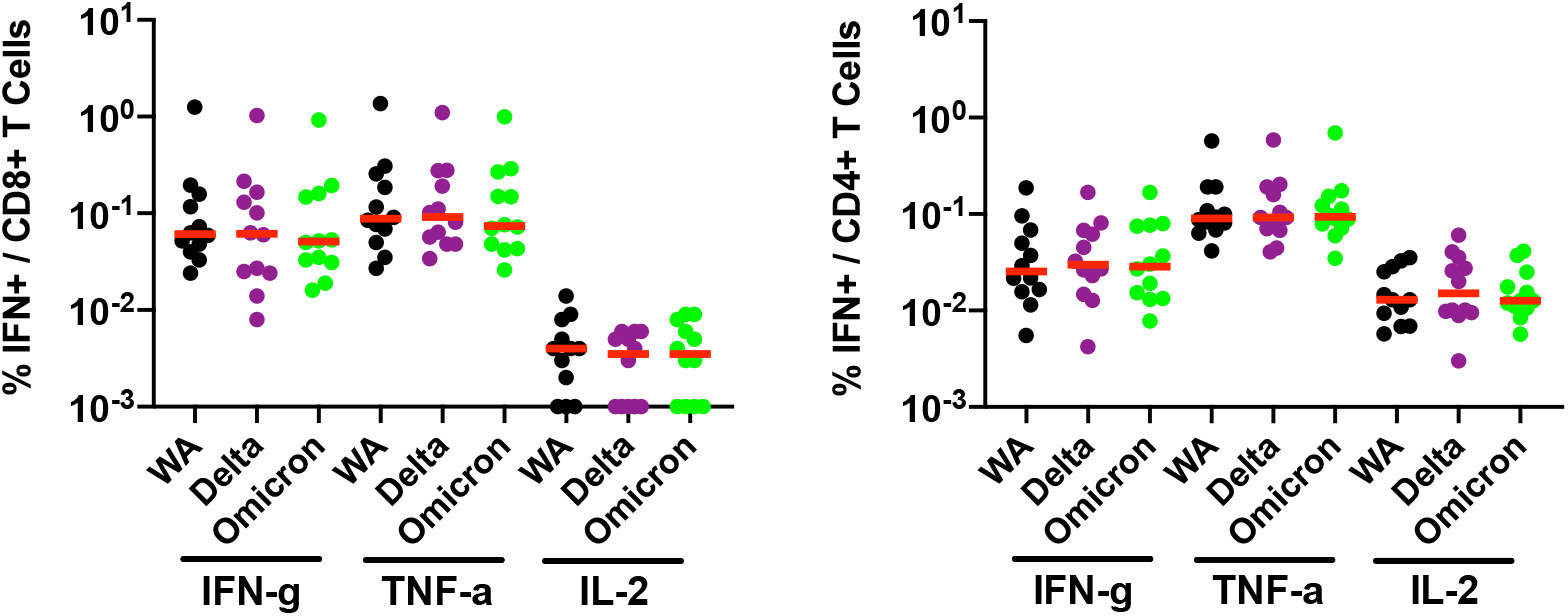
Cellular immune responses to Omicron by intracellular cytokine staining assays. Spike-specific IFN-γ, TNF-*α*, and IL-2 CD8+ and CD4+ T cell responses by intracellular cytokine staining assays at month 8 following vaccination with Ad26.COV2.S. Responses were measured against the SARS-CoV-2 WA1/2020, B.1.617.2 (Delta), and B.1.1.529 (Omicron) variants. Medians (red bars) are depicted and numerically shown.

